# Frailty and rate of fractures in patients initiating antihypertensive medications: a cohort study in primary care

**DOI:** 10.1101/2021.10.02.21264455

**Authors:** Marc F Österdahl, Sarah-Jo Sinnott, Ian Douglas, Andrew Clegg, Laurie Tomlinson, Angel Wong

## Abstract

**Background:** Treatment for hypertension improves cardiovascular outcomes. Frailty is common in people treated for hypertension, and associated with increases in adverse drug effects, potentially including falls resulting in fractures. We aimed to determine the association between baseline frailty and fractures in patients initiated on antihypertensive treatment.

**Methods:** We conducted a retrospective cohort study using United Kingdom primary care data, including new-users of first-line antihypertensives aged 65 years or over. We reported degree of frailty (fit, mild, moderate, severe) at antihypertensive initiation using the Electronic Frailty Index. We examined the association of frailty with fractures using multivariable Poisson regression, and assessed for interaction between antihypertensive class and frailty.

**Results:** 49634 (43%) people initiated on first-line antihypertensives were mildly or more frail. Over 4.1 years mean follow-up, 6567 (5.8%) experienced a fracture, with 3832 (58%) of these fractures occurring in frail people. Among those with severe frailty doubling of fracture risk was observed after antihypertensive initiation, compared with fit people [adjusted rate ratio 2.26 (95% CI 1.93-2.65)]. This pattern was replicated for hip and arm fractures, and strongest for spine fractures. The association between different types of antihypertensives and fractures varied by frailty (P=0.004), with a lower rate in moderately frail users of renin-angiotensin blockers compared with calcium-channel blockers (RR 0.81 95% CI 0.71-0.94)

**Conclusions:** Frailty is common among people initiating first-line antihypertensive treatment, and was associated with an increased fracture rate. Awareness of this is important to encourage clinicians to consider risk of falls and fractures when treating hypertension.

## 1. Introduction

Hypertension is the 4th most common risk factor driving mortality and morbidity in the UK[1] and is more common with increasing age[2]. Recent evidence from randomised controlled trials shows benefit from drug treatment in older adults, and possibly older people with frailty[3][4][5]. However, there are concerns about adverse effects of antihypertensive medications, including syncope and possibly falls[6][7], leading to fractures especially amongst elderly and frail patients. Such fractures can occur with minimal trauma, such as a fall from standing height, and are labelled ‘fragility fractures’[8]. These place a substantial burden on countries’ health systems, both due to immediate care costs and consequent long-term disability. Hip fractures are associated with particularly high mortality and morbidity[9], representing 54% of the €37 billion cost of all fractures in the European Union by 2010[10].

Age alone is increasingly recognised as less useful than frailty for predicting many outcomes experienced by older patients, including mortality and fractures[11][12]. Frailty is a condition characterised by loss of biological reserves across multiple organ systems and vulnerability to physiological decompensation after a stressor event[13]. It is distinct from both co-morbidity and disability, although incorporates elements of both, and may be partly reversible[14][15]. Frailty can be quantified using the electronic frailty index (eFI) [13], created using data from routine General Practice consultations in the UK and based on the well-established cumulative deficit model of frailty[16]. It is now used both for research and recommended by NICE for use in clinical practice[17][18].

The evidence regarding the impact of antihypertensive treatment on fracture risk is mixed. There is observational evidence of an increase in risk for the first 45 days[19]. The recent HYpertension in the Very Elderly Trial (HYVET) showed possible reductions in fracture risk for active treatment compared to placebo, but in a secondary analysis, with a shorter follow-up time than originally planned. [20] [21] HYVET did not report the effects of different types of anti-hypertensive initiated on fracture risk [21]. However, ALLHAT (Antihypertensive and Lipid-Lowering Treatment to Prevent Heart Attack Trial), suggested a lower fracture risk for treatment with thiazide-type diuretics compared to ACE/ARB, but not calcium-channel blockers [22].

Awareness of an association between frailty and adverse outcomes, such as fractures, would help inform clinicians on the risks and benefits of initiating antihypertensive treatment in people with different levels of frailty. We therefore sought to examine the association between frailty, identified using the eFI, and the rate of fractures in people initiating antihypertensive therapy in contemporary UK primary care and whether this varied by type of antihypertensive initiated.

## 2. Methods

### 2.1 Study Design

#### 2.1.1 Data Source

This study was a population-based cohort extracted from the Clinical Practice Research Datalink-Gold (CPRD) database for a previous study[23]. This database contains anonymised primary care data for approximately 7% of the UK population, submitted by enrolled General Practitioners (GPs). It is considered representative of the UK population in terms of age, sex and ethnicity[24]. It includes coded information on demographics, diagnoses, symptoms & signs, as well as prescribing data and test results[25].

#### 2.1.2 Participants

We included adults enrolled in CPRD-Gold between 1 Jan 2007 and 31 December 2017, who started a National Institute for Clinical Excellence (NICE) recommended first-line antihypertensive medication. This could be either an angiotensin converting enzyme-inhibitor, or an angiotensin-receptor blocker (ACE/ARB), a calcium channel blocker (CCB), or a thiazide/thiazide-like diuretic[2,26]. Beta-blockers, alpha-blockers and loop diuretics were not recommended first line so were not included. As our exposure, eFI, is only validated for people aged 65 years or over, we excluded people aged less than 65[13]. To maximise the number of eligible participants we did not use linkage to secondary care data since our outcome, fracture, is already well-recorded in primary care records.[27][28] People were eligible for study inclusion from the latest date of: 01 January 2007, their 65th birthday, or 1 year after the participant’s registration in CPRD.

To ensure that the medication was started for hypertension rather than another indication, we required a blood pressure reading of over 140/90mmHg to have been recorded at starting date or within 1 year prior. We excluded participants commencing more than one drug simultaneously, or who had been prescribed any antihypertensive previously before study start date. (Figure 1)

**Fig. 1.**
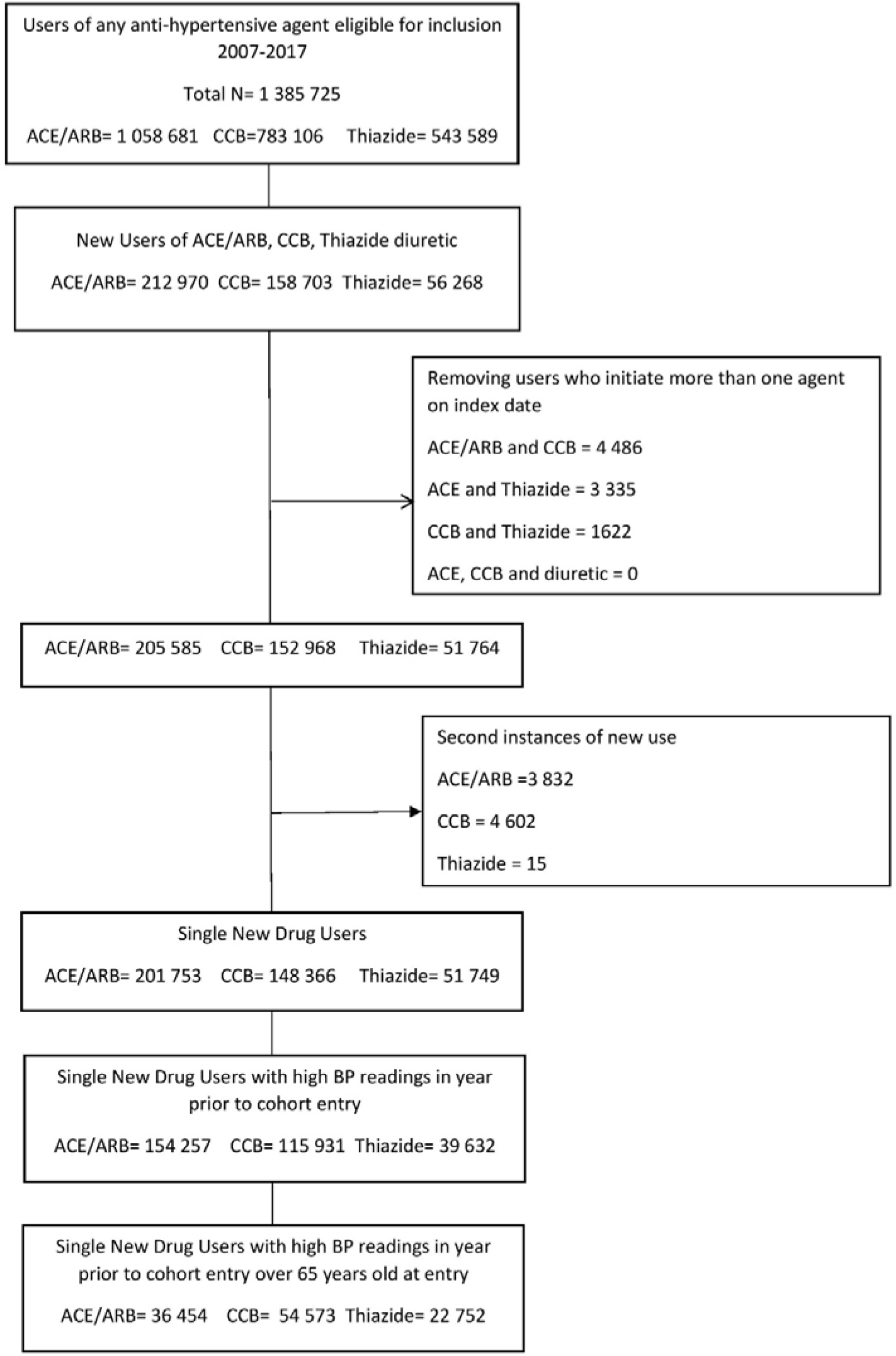
Study Flowchart ACE/ARB= Angiotensin Converting Enzyme Inhibitor or Angiotensin II receptor blocker, CCB= Calcium Channel blocker, Thiazide= Thiazide or Thiazide-like diuretic. BP = Blood Pressure

#### 2.1.3 Exposure

We defined frailty using the electronic frailty index (eFI), with the addition of terminal illness [13]. This uses a cumulative deficit model of frailty, counting 37 components associated with ageing and adverse outcomes, to generate a score. A component is present if a relevant abnormal test result, clinic measurement or Read code is associated with the patient at the time of antihypertensive drug initiation. We used pre-established cut points from the original eFI development cohort, to define four categories of frailty: “Fit” (score<=0.12), “Mild Frailty” (score 0.12-0.24), “Moderate Frailty” (score 0.24-0.36) and “Severe Frailty” (score >0.36).

#### 2.1.4 Outcome

Our primary outcome was any fragility fracture. This was extracted from CPRD, using pre-established Read codes lists for hip, arm, or spine fractures. Previous work has validated the recording of fractures in CPRD[27]. Sites were chosen in line with those included in NICE guidelines on Osteoporosis, and previous studies using CPRD[8,29]. An additional category of fragility fractures of unspecified location was also used. Individual fracture sites were analysed as secondary outcomes. We followed participants from the date of the initiation of antihypertensives until the earliest of: occurrence of their first fracture, death, or deregistration from a CPRD practice.

#### 2.1.5 Covariates

We defined Covariates a priori based on previous knowledge and review of the relevant literature [8][30,31]. We included biological covariates of age and sex, and lifestyle covariates of smoking, alcohol consumption and body-mass-index (BMI), osteoporosis (defined as the presence of a Read code for osteoporosis or a previous fragility fracture), calendar year (to account for changing guidelines regarding treatment of hypertension [32][26]), and seasonal effect of injury (winter/summer) (Figure 1). (**Supplementary Figure 1)**.

#### 2.1.6 Missing Data

Low levels of missing data (<5%) were expected for smoking and alcohol consumption, and for this dataset this was also known to be the case for BMI [23] The selected co-variates are known to be acceptably recorded in CPRD, and low levels of missing data were expected[24,33]. We therefore used complete case analysis in our fully adjusted model. We anticipated more missing data for ethnicity, known to be poorly recorded in CPRD[34][35], so confined this to sensitivity analysis. Deprivation data is only available through linkage, which is only possible for 60% of practices[24]. The use of bisphosphonates is only available as prescriptions issued, but as treatment compliance is known to be poor this was considered insufficient for inclusion in primary analysis[36]. The same is true for calcium and vitamin D supplementation, although their effect on fractures is less marked[37,38].

### 2.2. Statistical Analysis

We conducted Poisson regression to calculate rate ratios (RR) and 95% confidence intervals (95% CI) for any fracture according to frailty category with ‘Fit’ as the reference group. We conducted a univariable model, an age- and sex-adjusted model, and a multivariable fully-adjusted model. Our fully-adjusted model assessing association between frailty and fracture was adjusted for age, sex, BMI, smoking, alcohol, osteoporosis, season and year of study. All data management and analyses were performed using Stata 16 (StataCorp, Texas).

### 2.2.1 Secondary & Sensitivity Analysis

In a pre-planned secondary analysis, we calculated rate of fracture for each fracture site, adjusting for the same co-variates as the primary analysis. We pre-specified analysis of an interaction between frailty and the class of antihypertensive on the risk of fracture, using a likelihood ratio test for interaction. In sensitivity analyses, due to anticipated missing data, we also adjusted separately for ethnicity and patient-level index of multiple deprivation[24,25]. In addition, we also explored the effect of bisphosphonate as a potential confounder of the model, although compliance is known to be poor compared to prescription.

## 3. Results

### 3.1 Study Population and baseline Characteristics

We identified 113,779 people aged 65 or over who initiated either ACEI/ARB, CCB or thiazide diuretic between 2007-2017. Mean follow-up was 4.1 years (standard deviation (SD) 2.8 years), yielding a total of 466,923 person-years of follow-up (**Figure 1**).

A total of 32% (36,454/113,779) initiated ACEI/ARB, 48% (54,573/113,779) a CCB and 20.0% (22,752/113,779) thiazide diuretics. The proportions changed with age, with 34.7% (13,329/38,438) of patients aged 65-69 years initiating ACEI/ARB, compared to 28.0% (2,632/9,405) aged over 85 years, whilst 15.7% (6,045/38,438) of those aged 65-69 years were prescribed thiazide diuretics, compared to 28.9% (2,714/9,405) aged over 85 years. The proportion of people initiating thiazide diuretics fell each year from 28.5% (4,916/17,221) in 2007 to 6.4% (275/4,313) in 2017, with CCB use progressively increasing from 32.7% (5,632/17,221) in 2007 to 70.9% (3,056/4,313) in 2017. (**Supplementary Table 1**).

### 3.2 Frailty

The majority of participants (56.4% (64,145/113,779)) were classified as ‘fit’, 32.0% (36,373/113,779) as ‘mild frailty’, 10.5% (11,904/113,779) ‘moderate frailty’ and 1.2% (1357/113,779) ‘severe frailty’ (**Table 1**). Older people were more likely to be categorised as frail: participants who were ‘fit’ had a mean age of 71 years (SD 6) compared to 82 years (SD 8) for people categorised as ‘severely frail’. People categorised as ‘severely frail’ were more likely to be underweight than the ‘fit’ (6.1% vs 1.3%), less likely to smoke and drink alcohol, more likely to have a diagnosis of osteoporosis (57.9% vs 7.8%), to have been prescribed a bisphosphonate (44.9% vs 7.8%) and to be the most deprived (10.5% vs 6.0%).

**Table 1:**
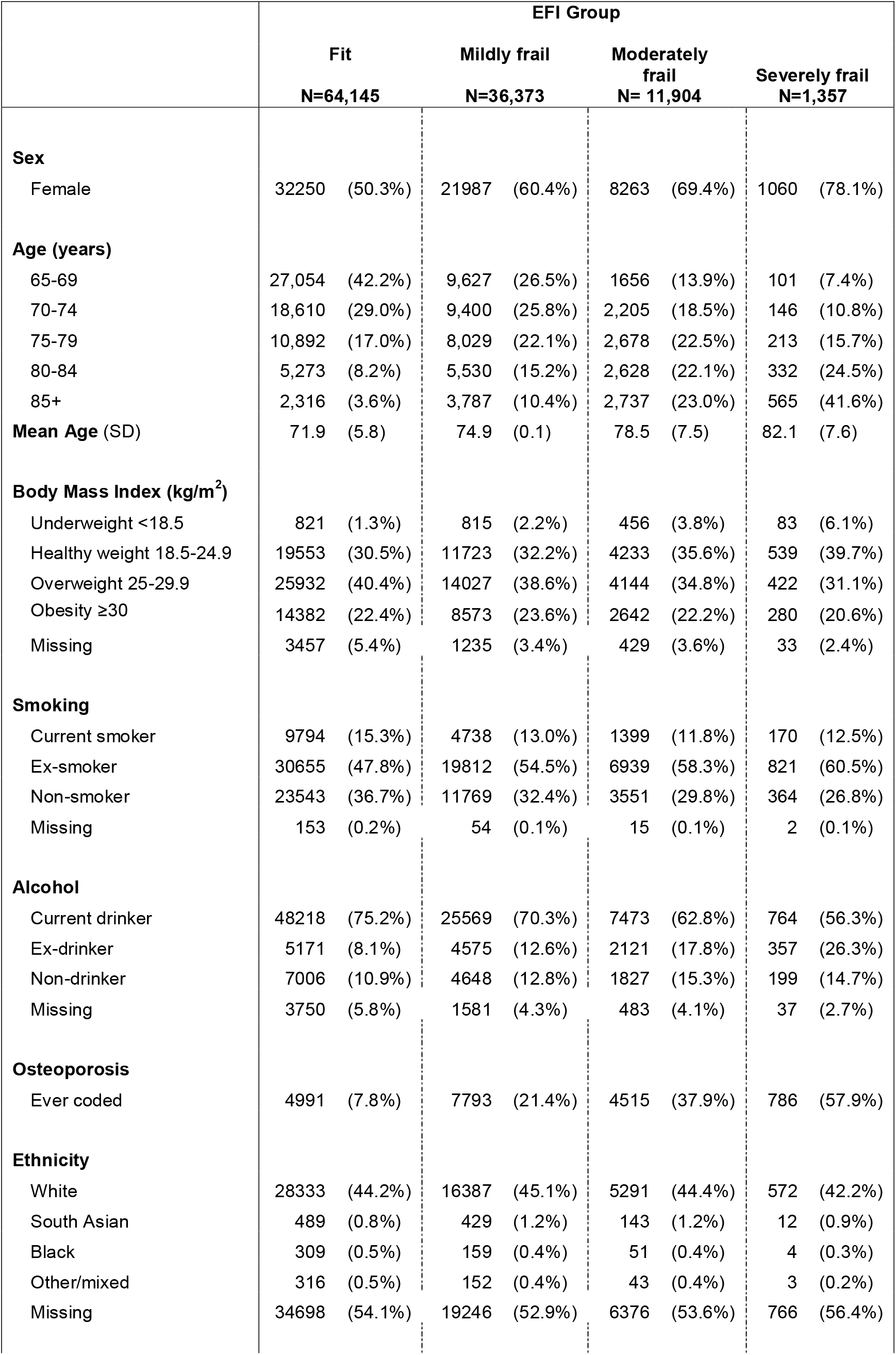
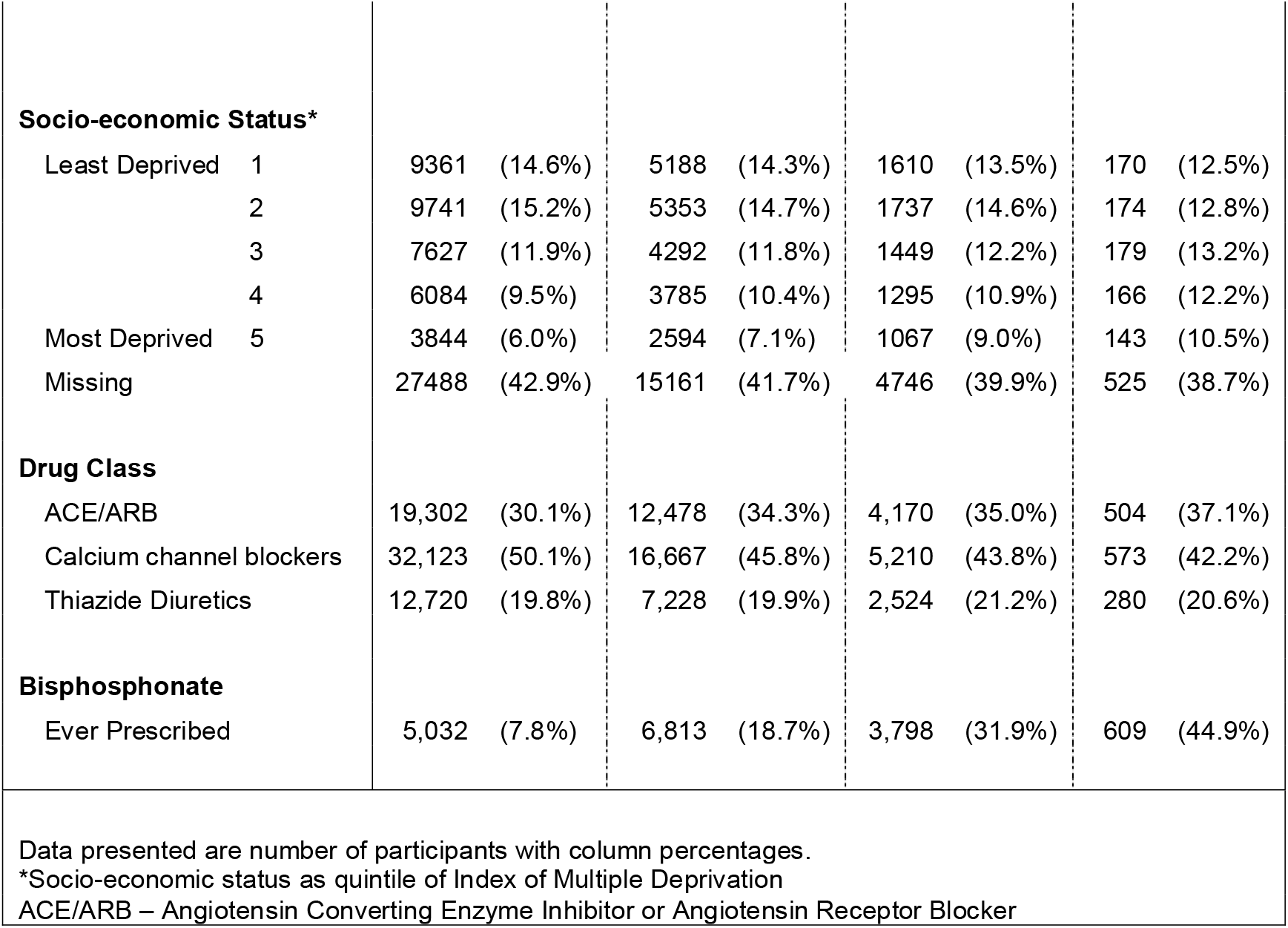
Associations between eFI group and baseline characteristics of cohort.

### 3.3 Fractures

6,567 (5.8%) experienced a fracture during follow-up. This yielded an unadjusted fracture rate of 14.1/1000 person years at risk (PYAR). Fracture rate increased with increasing age and female sex, as well as low BMI and osteoporosis (**Supplementary Tables 2 and 3**). Hip fractures were 32% of all recorded first fractures, whilst 35% were arm fractures, 13% spinal and 20% were fragility fractures without a specified site. (**Table 2**)

**Table 2:** Rate ratio for any fracture in each frailty category for each drug class.

| Drug Class | eFI Group |  |  |  |
| --- | --- | --- | --- | --- |
|  | Fit | Mild | Mod | Severe |
| ACE/ARB | 1<br>(ref) | 1.53<br>(1.38-1.70) | 1.68<br>(1.46-1.92) | 2.73<br>(2.12-3.51) |
| CCB | 1<br>(ref) | 1.37<br>(1.26-1.50) | 1.91<br>(1.71-2.14) | 2.58<br>(2.03-3.28) |
| Thiazide | 1<br>(ref) | 1.20<br>(1.07-1.34) | 1.62<br>(1.41-1.87) | 1.76<br>(1.25-2.48) |
| Summary | 1<br>(ref) | 1.37<br>(1.29-1.45) | 1.75<br>(1.62-1.89) | 2.40<br>(2.05-2.82) |

### 3.4 Association of frailty with Rate of Fractures

58% of all fractures, and 63% of hip fractures occurred in patients with some degree of frailty (**Figure 2)**. In the fully adjusted model, people with severe frailty experienced an increased rate of any fracture compared to those in the fit category (RR 2.26, 95% CI 1.93-2.65). An increased rate was also seen for those categorised with moderate frailty (RR 1.68, 95% CI 1.56-1.82) and mild frailty (RR1.34, 95% CI 1.26-1.43), compared to the fit category. This association between frailty and fracture rate remained for all fracture sites, but strongest for spine (RR 3.49, 95% CI 2.31-5.25) and weakest for arm (RR1.95, 95% CI 1.46-2.62) **(Figure 3, Supplementary Table 4)**

**Fig. 2.**
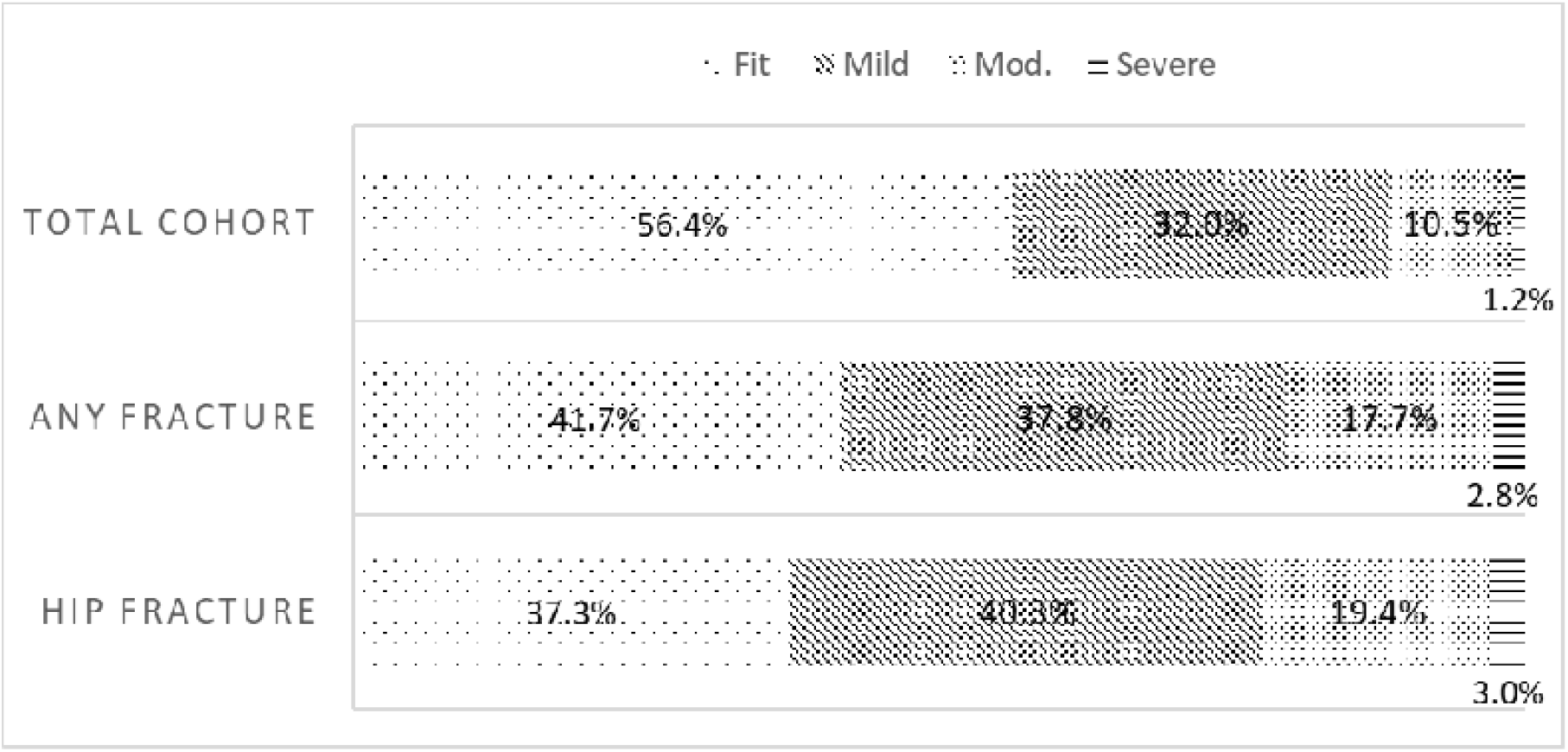
Proportion of: Total cohort in each eFI group; those experiencing any fracture by eFI group; experiencing hip fractures by eFI group

**Fig. 3.**
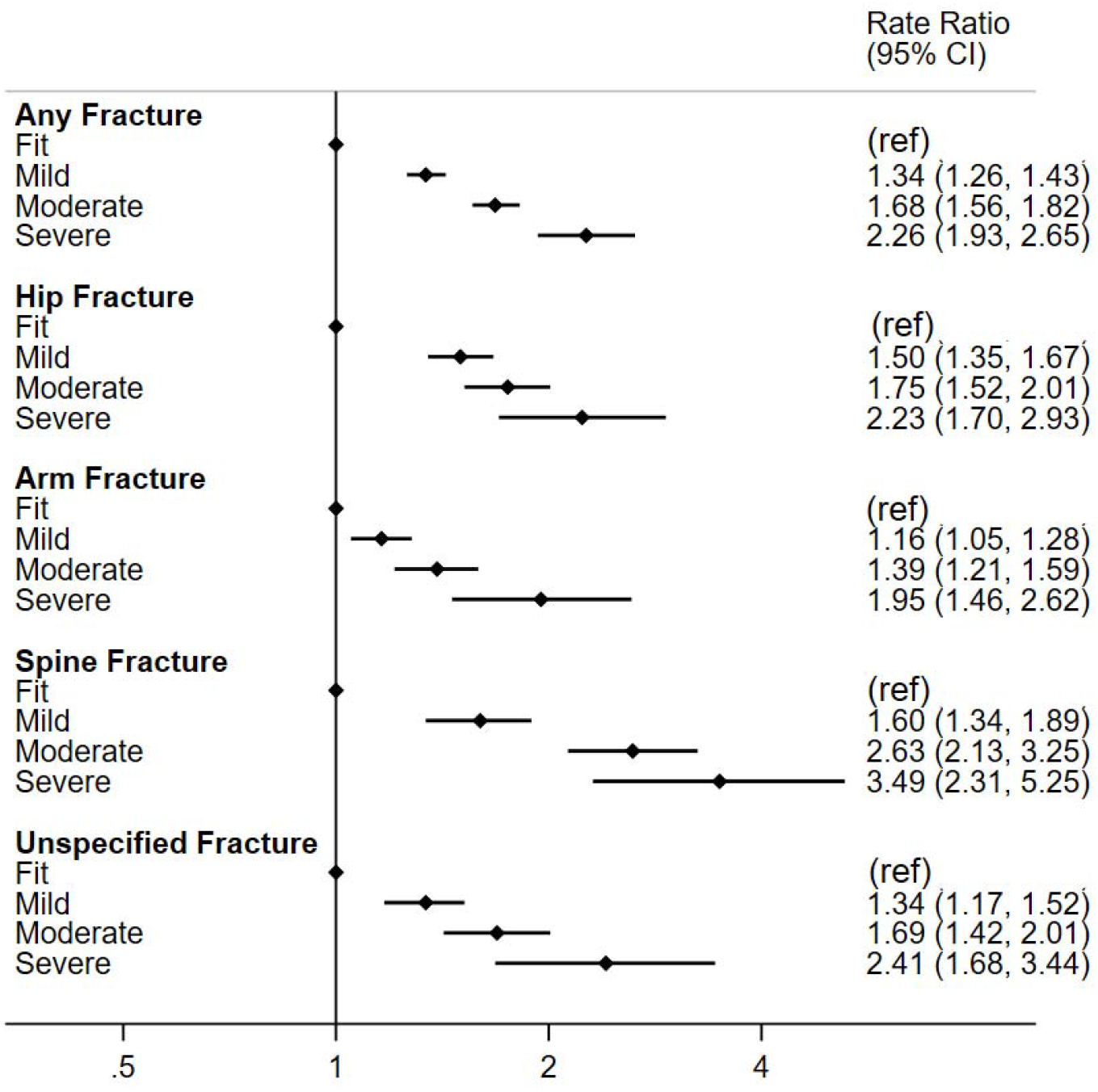
Fully Adjusted Rate ratio for Fracture at each site by eFI category Adjusted for age, sex, BMI, smoking, alcohol, osteoporosis, season and year of study

### 3.5 Sensitivity Analyses

We did not see any meaningful difference from our estimates for the primary analysis for any of the sensitivity analyses (**Supplementary Table 5** and **Figure 4**).

**Fig. 4.**
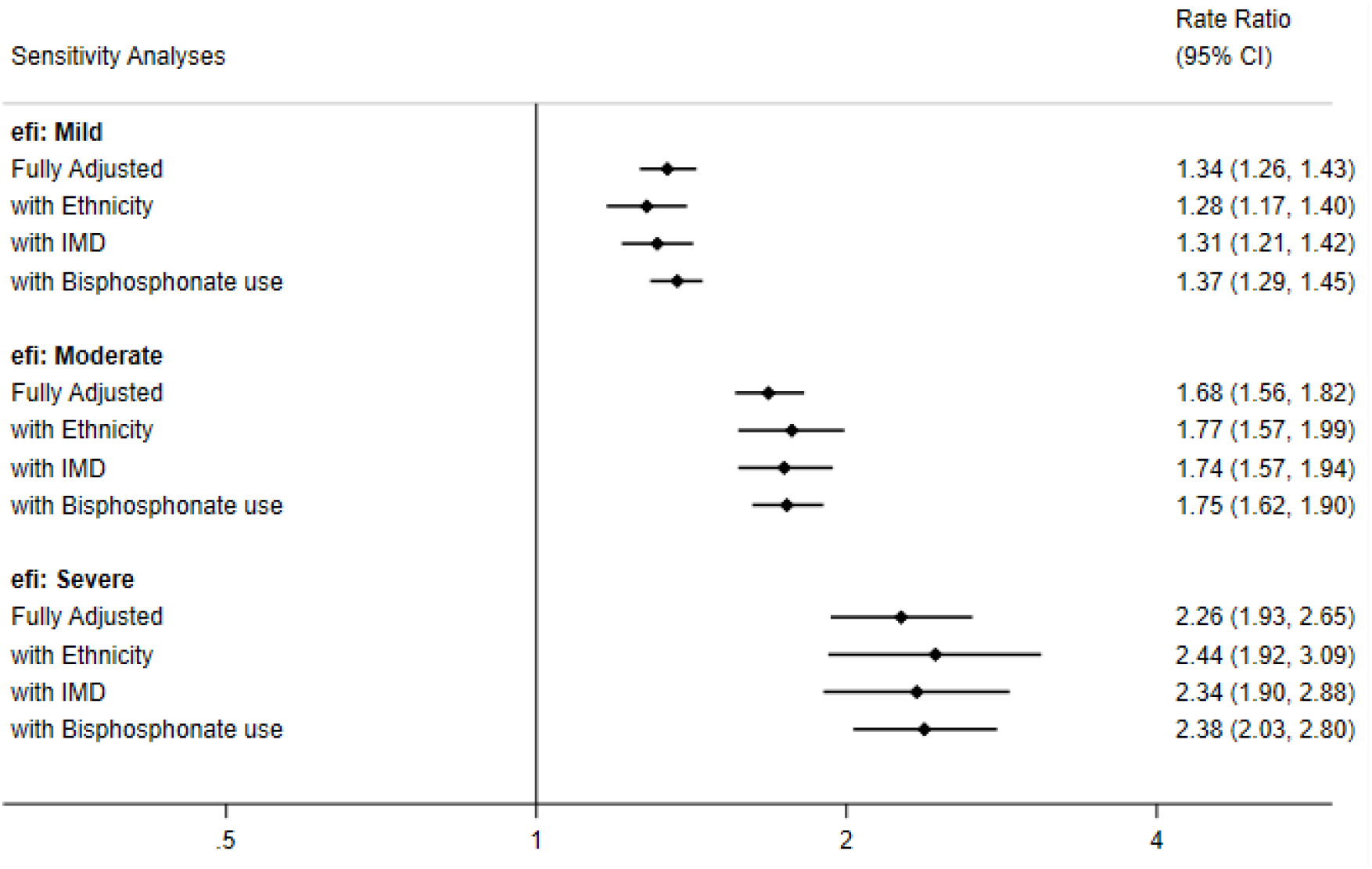
Sensitivity Analyses on rate of any fracture by eFI Group Additionally adjusted for age, sex, osteoporosis season, year of study, smoking, alcohol and BMI

### 3.6 Fracture rate by drug class

Increasing frailty increased the rate of fractures for all drug classes (**Table 2**). There was no evidence for a difference in fracture rate by drug class without accounting for eFI (**Supplementary Table 6**)

We found strong evidence that the association between type of antihypertensives and rate of fracture varied by frailty (p=0.004). In particular, ACE/ARB users with moderate frailty had a lower rate of fracture than moderately frail Calcium Channel Blocker users (RR 0.81 95% CI 0.71-0.94) (**Table 3**)

**Table 3:** Rate ratio for any fracture for each drug class by eFI Group - CCB as reference.

| Drug Class | eFI Group |  |  |  | Summary Adjusted RR |
| --- | --- | --- | --- | --- | --- |
|  | Fit | Mild | Mod | Severe |  |
| CCB | 1.00<br>- | 1.00<br>- | 1.00<br>- | 1.00<br>- | 1.00<br>- |
| ACE/ARB | 0.93<br>(0.84-1.02) | 1.03<br>(0.94-1.14) | 0.81<br>(0.71-0.94) | 0.98<br>(0.71-1.36) | 0.95<br>(0.89-1.01) |
| Thiazide | 1.07<br>(0.98-1.18) | 0.94<br>(0.84-1.04) | 0.91<br>(0.79-1.06) | 0.73<br>(0.49-1.10) | 0.98<br>(0.92-1.05) |

## 4. Discussion

In this large cohort study reflecting routine clinical care we have demonstrated an increased risk of fractures among people initiating antihypertensives associated with every level of increased frailty relative to fit people. We also found evidence that association between type of antihypertensives and fractures varied by frailty, with a lower risk of fractures associated with ACE/ARB medications, compared with calcium channel blockers among people with moderately frailty.

The overall association between increasing frailty, as defined by eFI, and increasing rates of fracture is in line with our hypothesis and current literature[12,29]. The association between fracture site and eFI was strongest for spinal fractures, and weakest for arm. This may be because arm fractures may occur due to high impact injuries sustained by physically fit and active participants while spinal fractures in this group are often fragility fractures. This suggests that eFI is particularly helpful to clinicians, as fragility fractures are of great concern, but may be preventable through implementation of evidence-based interventions[8].

We observed a lower risk of fractures associated with ACE/ARB than CCB in moderately frail people but this requires further work in other cohorts to see if the association can be replicated. If causal, it would suggest calcium-channel blockers are less suited to moderately frail participants. It is possible that some of the association between CCB users and increased fracture rate, compared with ACE/ARB is due to residual confounding, if there are deficits related to frailty not captured by eFI, and ACE/ARBs were preferentially prescribed to less frail participants. However, a recent secondary analysis of the ALLHAT trial has suggested that CCBs increased the risk of falls in the first year of treatment, compared to an ACE or Thiazide[39]. This could lead to fractures, but fractures were not assessed by the trial. However, this was not sustained over the entire follow-up period, nor did they stratify by frailty. Ideally association between fracture risk and class of antihypertensive would be compared prospectively in a trial, although as seen by HYVET, large numbers would be needed to have power to compare different treatments for hypertension. For clinicians, the relationship between frailty and fractures, and potentially drug class, is an important relationship to be aware of, so that they might take further measures to assess a frail person’s risk of fracture when starting an antihypertensive. This can help trigger shared decision making between clinicians and frailer patients, allowing them to benefit both from the potential reduction in cardiovascular risk, whilst mitigating their increased fracture risk.

We have conducted a large cohort study, with long follow-up time and a wide distribution of age which was well-powered for each level of frailty and able to assess site-specific fractures. Our results are reflective of treatment patterns and outcome rates in routine clinical care. Our outcome of fracture is clinically objective and well recorded in CPRD[27]. We were able to adjust for detailed covariates, including those involved in the eFI with little (<4%) missing data.

However, there were also limitations, in particular that despite detailed covariate adjustment there could have been residual confounding. Although results were not affected by adjustment for ethnicity, deprivation, and bisphosphonate use in sensitivity analyses, data on steroid use, calcium supplementation, or vitamin D levels were not available. These factors could be linked to frailty and fracture rate[8,31]. Osteoporosis is likely to be underdiagnosed in this population as it is asymptomatic before the first fracture[40]. Although there is some data on falls in CPRD, used to calculate eFI, it is not validated and does not include information on frequency of falls, and likely under-reported [41]. Although the eFI is now widely used clinically and in research, it relies on the assumption that the absence of a Read code, or abnormal result, indicates the absence of the deficit. There has been limited validation of CPRD records for some deficits, which may have underestimated the true prevalence and severity of frailty in our population, and bias our estimate towards null. Conversely, codes may remain on a patient’s record, (e.g. following a period of being housebound) even if no longer relevant.. Despite these limitations, eFI has been successfully validated by comparison with other internationally established measures of frailty [42].

Finally, CPRD also only provides data on prescriptions issued which may not reflect actual medication use by participants. Antihypertensive adherence can be poor and may be worse with increased frailty[43] but this would likely bias our findings to the null.

## 5. Conclusion

We demonstrate that a substantial proportion of people initiating antihypertensive treatment are frail, and that frailty is associated with subsequent rate of fractures. We have also demonstrated that the association between different classes of antihypertensive and fractures varied by frailty with an increased risk of fractures associated with calcium channel blockers compared with ACE/ARB in moderately frail people. Clinicians and patients should be aware of these associations to weigh the potential benefits of antihypertensive therapy against the risk of fractures in the context of frailty.

## Data Availability

The datasets supporting this work are not publicly available due to CPRD licencing restrictions. Codelists for variables for fractures are however available through LSHTM data repository.

## 6. Ethical Declarations

### Code availability

Codelists for variables for fractures are however available through LSHTM data repository, and more detailed code available upon request from authors.

### Author’s Contributions

MFÖ, LT and AW, conceived the study in conjunction with SJS, ID and AC. AC developed the electronic frailty index. MFÖ conducted the analysis and wrote the first draft of this manuscript. All authors reviewed the manuscript, contributed to the interpretation of results and to its final form.

### Ethics Approval

Ethical approval was granted for this project by the London School of Hygiene and Tropical Medicine Research Ethics committee (Ref: 16450) and by the Independent Scientific Advisory Committee (ISAC) of CPRD as a subset of a larger project (ISAC Protocol Number: 18_312R).

