## Supplementary Material for "Frailty and rate of fractures in patients initiating antihypertensive medications: a cohort study in primary care"

**Journal:** Drugs & Therapy Perspectives

**Appendix 1: Components of the Electronic Frailty Index**

| Deficit | Prevalence |
|---|---|
| **Anaemia and haematinic deficiency** | 10.6% |
| **Arthritis** | 20.6% |
| **Atrial Fibrillation** | 1.7% |
| **Cerebrovascular Disease** | 4.2% |
| **Chronic Kidney Disease** | 9.1% |
| **Diabetes** | 9.7% |
| **Dizziness** | 18.2% |
| **Dyspnoea** | 12.1% |
| **Falls** | 8.4% |
| **Foot Problems** | 2.6% |
| **Fragility Fracture** | 6.2% |
| **Hearing Impairment** | 11.8% |
| **Heart Failure** | 0.6% |
| **Heart Valve Disease** | 0.6% |
| **Housebound** | 12.0% |
| **Hypertension** | 21.6% |
| **Hypotension/Syncope** | 6.8% |
| **Ischaemic Heart Disease** | 21.2% |
| **Limitation in Physical Activity** | 0.6% |
| **Memory and cognitive problems** | 2.3% |
| **Mobility and transfer problems** | 1.3% |
| **Osteoporosis** | 4.8% |
| **Parkinsonism and tremor** | 0.7% |
| **Peptic Ulcer** | 3.9% |
| **Peripheral Vascular Disease** | 1.3% |
| **Polypharmacy** | 37.4% |
| **Requirement for Care** | 0.6% |
| **Respiratory Disease** | 23.2% |
| **Skin Ulcer** | 7.7% |
| **Sleep disturbance** | 9.5% |
| **Social Vulnerability** | 2.1% |
| ***Terminal Disease** | 0.3% |
| **Thyroid Disease** | 9.2% |
| **Urinary Incontinence** | 4.6% |
| **Urinary System disease** | 26.0% |
| **Visual Impairment** | 11.3% |
| **Weight loss and anorexia** | 3.4% |

*Terminal disease was not included in the original eFI in 2016

### Appendix 2: Supplementary Tables + Figures

| | ACE/ARB | | CCB | | Thiazide | | Total |
|---|---|---|---|---|---|---|---|
| **Total** | 36,454 | (32.0%) | 54,573 | (48.0%) | 22,752 | (20.0%) | 113,779 |
| **Sex** | | | | | | | |
| Male | 18,565 | (37.0%) | 24,034 | (47.9%) | 7,620 | (15.2%) | 50,219 |
| Female | 17,889 | (28.1%) | 30,539 | (48.0%) | 15,132 | (23.8%) | 63,560 |
| **Age** | | | | | | | |
| 65-69 | 13,329 | (34.7%) | 19,064 | (49.6%) | 6,045 | (15.7%) | 38,438 |
| 70-74 | 9,722 | (32.0%) | 14,852 | (48.9%) | 5,787 | (19.1%) | 30,361 |
| 75-79 | 6,667 | (30.6%) | 10,367 | (47.5%) | 4,778 | (21.9%) | 21,812 |
| 80-84 | 4,104 | (29.8%) | 6,231 | (45.3%) | 3,428 | (24.9%) | 13,763 |
| 85+ | 2,632 | (28.0%) | 4,059 | (43.2%) | 2,714 | (28.9%) | 9,405 |
| **Mean Age** | 73.2 | | 73.4 | | 75.1 | | 73.7 |
| (standard deviation) | (6.65) | | (6.66) | | (7.19) | | (6.81) |
| **BMI** | | | | | | | |
| underweight <18.5 | 599 | (27.5%) | 1,069 | (49.1%) | 507 | (23.3%) | 2,175 |
| healthy weight 18.5-24.9 | 10,697 | (29.7%) | 17,696 | (49.1%) | 7,655 | (21.2%) | 36,048 |
| overweight 25-29.9 | 14,579 | (32.7%) | 21,573 | (48.5%) | 8,373 | (18.8%) | 44,525 |
| obesity >=30 | 9,107 | (35.2%) | 11,688 | (45.2%) | 5,082 | (19.6%) | 25,877 |
| Missing | 1,472 | (28.6%) | 2,547 | (49.4%) | 1,135 | (22.0%) | 5,154 |
| **Smoking** | | | | | | | |
| Current smoker | 5,237 | (32.5%) | 7,742 | (48.1%) | 3,122 | (19.4%) | 16,101 |
| Ex-smoker | 19,316 | (33.2%) | 27,815 | (47.8%) | 11,096 | (19.1%) | 58,227 |
| Non-smoker | 11,859 | (30.2%) | 18,894 | (48.2%) | 8,474 | (21.6%) | 39,227 |
| Missing | 42 | (18.8%) | 122 | (54.5%) | 60 | (26.8%) | 224 |
| **Alcohol** | | | | | | | |
| Current drinker | 26,289 | (32.1%) | 39,809 | (48.5%) | 15,926 | (19.4%) | 82,024 |
| Ex-drinker | 4,058 | (33.2%) | 5,794 | (47.4%) | 2,372 | (19.4%) | 12,224 |
| Non-drinker | 4,274 | (31.2%) | 6,178 | (45.2%) | 3,228 | (23.6%) | 13,680 |
| Missing | 1,833 | (31.3%) | 2,792 | (47.7%) | 1,226 | (21.0%) | 5,851 |
| **Osteoporosis** | | | | | | | |
| Yes | 5055 | (28.0%) | 8888 | (49.1%) | 4142 | (22.9%) | 18,085 |
| **Ethnicity** | | | | | | | |
| white | 16376 | (32.4%) | 24659 | (48.7%) | 8548 | (16.9%) | 50,583 |
| South Asian | 431 | (40.2%) | 510 | (47.5%) | 132 | (12.3%) | 1,073 |
| Black | 123 | (23.5%) | 334 | (63.9%) | 66 | (12.6%) | 523 |
| other/mixed | 157 | (30.5%) | 273 | (53.1%) | 84 | (16.3%) | 514 |
| Missing | 19367 | (31.7%) | 28797 | (47.1%) | 22752 | (37.2%) | 61,086 |
| **Deprivation (IMD Quintile)** | | | | | | | |
| Least Deprived - 1 | 5,411 | (33.1%) | 7,961 | (48.8%) | 2,957 | (18.1%) | 16,329 |
| 2 | 5,642 | (33.2%) | 8,136 | (47.8%) | 3,227 | (19.0%) | 17,005 |
| 3 | 4,553 | (33.6%) | 6,391 | (47.2%) | 2,603 | (19.2%) | 13,547 |
| 4 | 3,717 | (32.8%) | 5,376 | (47.4%) | 2,237 | (19.7%) | 11,330 |

| | | | | | | | |
|---|---|---|---|---|---|---|---|
| Most Deprived - 5 | 2,444 | (32.0%) | 3,607 | (47.2%) | 1,597 | (20.9%) | 7,648 |
| Missing | 14,687 | (30.6%) | 23,102 | (48.2%) | 10,131 | (21.1%) | 47,920 |
| **Bisphosphonate** | | | | | | | |
| Ever Prescribed | 4,754 | (29.3%) | 7,618 | (46.9%) | 3,880 | (23.9%) | 16252 |
| **Year of Drug Initiation** | | | | | | | |
| 2007 | 6,673 | (38.7%) | 5,632 | (32.7%) | 4916 | (28.5%) | 17,221 |
| 2008 | 5,629 | (36.5%) | 5,473 | (35.5%) | 4324 | (28.0%) | 15,426 |
| 2009 | 4,947 | (35.9%) | 5,197 | (37.7%) | 3654 | (26.5%) | 13,798 |
| 2010 | 4,151 | (33.6%) | 5,072 | (41.0%) | 3141 | (25.4%) | 12,364 |
| 2011 | 3,396 | (31.7%) | 5,073 | (47.4%) | 2236 | (20.9%) | 10,705 |
| 2012 | 2,953 | (28.6%) | 6,027 | (58.4%) | 1342 | (13.0%) | 10,322 |
| 2013 | 2,684 | (27.9%) | 5,839 | (60.7%) | 1100 | (11.4%) | 9,623 |
| 2014 | 2,124 | (26.6%) | 5,067 | (63.5%) | 794 | (9.9%) | 7,985 |
| 2015 | 1,730 | (25.4%) | 4,515 | (66.3%) | 563 | (8.3%) | 6,808 |
| 2016 | 1,185 | (22.7%) | 3,622 | (69.5%) | 407 | (7.8%) | 5,214 |
| 2017 | 982 | (22.8%) | 3,056 | (70.9%) | 275 | (6.4%) | 4,313 |

**Supplementary Table 1: Baseline Characteristics by Antihypertensive Drug**
Data presented with row percentages
ACE/ARB = Angiotensin Converting Enzyme Inhibitor or Angiotensin-receptor blocker, CCB= Calcium Channel Blocker
BMI= Body Mass Index, IMD= Index of Multiple Deprivation

| | Total number | (Col%) | Person-Years | (Col%) | Fractures | Rate (/1000pyears) | Crude Rate Ratio | (95% CI) |
|---|---|---|---|---|---|---|---|---|
| **Total Cohort** | 113779 | | 466923 | | 6576 | 14.06 | | |
| **Sex** | | | | | | | | |
| Male | 50,219 | (44%) | 209557 | (45%) | 1295 | 6.18 | 1.00 | (baseline) |
| Female | 63,560 | (56%) | 257367 | (55%) | 5272 | 20.48 | 3.31 | (3.12-3.52) |
| **Age** | | | | | | | | |
| 65-69 | 38,438 | (77%) | 166091 | (79%) | 1326 | 7.98 | 1.00 | (baseline) |
| 70-74 | 30,361 | (60%) | 130060 | (62%) | 1500 | 11.53 | 1.44 | (1.34-1.56) |
| 75-79 | 21,812 | (43%) | 90494 | (43%) | 1530 | 16.91 | 2.12 | (1.97-2.28) |
| 80-84 | 13,763 | (27%) | 51830 | (25%) | 1228 | 23.69 | 2.97 | (2.75-3.21) |
| 85+ | 9,405 | (19%) | 28446 | (14%) | 983 | 34.56 | 4.33 | (3.99-4.70) |

**Supplementary Table 2: Unadjusted rate of any fracture in cohort**

| | Total number | (Col%) | Person-Years | (Col%) | Fractures | Adjusted Rate Ratio | (95% CI) |
|---|---|---|---|---|---|---|---|
| **Total Cohort** | 113779 | | 466923 | | 6576 | | |
| **BMI** | | | | | | | |
| underweight <18.5 | 2175 | (2%) | 6938 | (1%) | 264 | 1.63 | (1.45-1.83) |
| healthy weight 18.5-24.9 | 36048 | (32%) | 145145 | (31%) | 2756 | 1.00 | (baseline) |
| overweight 25-29.9 | 44525 | (39%) | 188991 | (40%) | 2234 | 0.77 | (0.73-0.80) |
| obesity >=30 | 25877 | (23%) | 109766 | (24%) | 1049 | 0.64 | (0.60-0.67) |
| Missing | 5154 | (5%) | 16082 | (3%) | 264 | | |

| | | | | | | | |
|---|---|---|---|---|---|---|---|
| **Smoking** | | | | | | | |
| Current smoker | 16101 | (14%) | 65525 | (14%) | 950 | 1.00 | (baseline) |
| Ex-smoker | 58227 | (51%) | 236045 | (51%) | 3083 | 0.81 | (0.75-0.87) |
| Non-smoker | 39227 | (34%) | 164635 | (35%) | 2520 | 0.76 | (0.71-0.82) |
| Missing | 224 | (0%) | 718 | (0%) | 14 | | |
| **Alcohol** | | | | | | | |
| Current drinker | 82024 | (72%) | 343175 | (73%) | 4420 | 1.00 | (baseline) |
| Ex-drinker | 12224 | (11%) | 46349 | (10%) | 807 | 1.09 | (1.01-1.18) |
| Non-drinker | 13680 | (12%) | 57887 | (12%) | 1017 | 0.99 | (0.92-1.06) |
| Missing | 5851 | (5%) | 19513 | (4%) | 323 | | |
| **Osteoporosis** | | | | | | | |
| Yes | 18085 | (16%) | 67006 | (14%) | 2057 | 1.86 | (1.76-1.96) |
| **Ethnicity** | | | | | | | |
| white | 50583 | (44%) | 206483 | (44%) | 2742 | 1.00 | (baseline) |
| South Asian | 1073 | (1%) | 3838 | (1%) | 25 | 0.54 | (0.36-0.80) |
| Black | 523 | (0%) | 2088 | (0%) | 9 | 0.37 | (0.19-0.72) |
| other/mixed | 514 | (0%) | 2022 | (0%) | 18 | 0.76 | (0.48-1.21) |
| Missing | 61086 | (54%) | 252492 | (54%) | 3773 | | |
| **Deprivation (IMD Category)** | | | | | | | |
| Least Deprived - 1 | 16329 | (14%) | 64492 | (14%) | 911 | 1.00 | (baseline) |
| 2 | 17005 | (15%) | 66289 | (14%) | 830 | 0.87 | (0.80-0.96) |
| 3 | 13547 | (12%) | 52259 | (11%) | 745 | 1.00 | (0.91-1.10) |
| 4 | 11330 | (10%) | 44548 | (10%) | 613 | 0.97 | (0.87-1.07) |
| Most Deprived - 5 | 7648 | (7%) | 29934 | (6%) | 421 | 1.00 | (0.89-1.12) |
| Missing | 47920 | (42%) | 209401 | (45%) | 3047 | | |
| **Bisphosphonate** | | | | | | | |
| Ever Prescribed | 16252 | (14%) | 69196 | (15%) | 1421 | 1.06 | (0.99-1.13) |
| **Participants contributing at least once to each season** | | | | | | | |
| Summer (May-Oct) | | | 235225 | (50%) | 3182 | 1.00 | (baseline) |
| Winter (Nov-April) | | | 231698 | (50%) | 3385 | 1.08 | (1.03-1.13) |
| **Participants contributing to each year of Study** | | | | | | | |
| 2007 | | | 8866 | (2%) | 97 | 1.00 | (baseline) |
| 2008 | | | 23662 | (5%) | 263 | 1.03 | (0.82-1.30) |
| 2009 | | | 35836 | (8%) | 504 | 1.33 | (1.07-1.65) |
| 2010 | | | 45070 | (10%) | 565 | 1.21 | (0.98-1.50) |
| 2011 | | | 51589 | (11%) | 653 | 1.24 | (1.00-1.54) |
| 2012 | | | 56777 | (12%) | 850 | 1.49 | (1.21-1.84) |
| 2013 | | | 58794 | (13%) | 887 | 1.54 | (1.25-1.89) |
| 2014 | | | 57261 | (12%) | 838 | 1.51 | (1.23-1.87) |
| 2015 | | | 51268 | (11%) | 768 | 1.57 | (1.27-1.94) |
| 2016 | | | 41684 | (9%) | 614 | 1.57 | (1.27-1.95) |
| 2017 | | | 36117 | (8%) | 528 | 1.59 | (1.28-1.97) |

**Supplementary Table 3 : Adjusted rate of any fracture by baseline characteristics of cohort**

Adjusted for Age + Sex only

Total number with column percentages

| | | | | EFI Group | | | |
|---|---|---|---|---|---|---|---|
| | | | **Total** | **Fit** | **Mild** | **Moderate** | **Severe** |
| | Participants | | 113779 | 64145 | 36373 | 11904 | 1357 |
| | PYAR | | 466,923 | 280104 | 143051 | 40200 | 3568 |
| **Any Fracture** | N (row %) | | 6,567 | 2735 (42) | 2485 (38) | 1165 (18) | 182 (3) |
| | Unadjusted Rate (/1000 PYAR) | | 14.1 | 9.8 | 17.4 | 29.0 | 51.0 |
| | Fully Adjusted | RR | | **1.00** | **1.34** | **1.68** | **2.26** |
| | | CI | | **(ref)** | (1.26-1.43) | (1.56-1.82) | (1.93-2.65) |
| **Hip Fracture** | N (row %) | | 2,080 | 775 (37) | 839 (40) | 403 (19) | 63 (3) |
| | Unadjusted Rate (/1000 PYAR) | | 4.5 | 2.8 | 5.9 | 10 | 17.7 |
| | Fully Adjusted | RR | | **1.00** | **1.50** | **1.75** | **2.23** |
| | | CI | | **(ref)** | (1.35-1.67) | (1.52-2.01) | (1.70-2.93) |
| **Arm Fracture** | N (row %) | | 2,325 | 1,108(48) | 824 (35) | 340(15) | 53(2) |
| | Unadjusted Rate (/1000 PYAR) | | 5.0 | 4.0 | 5.8 | 8.5 | 14.9 |
| | Fully Adjusted | RR | | **1.00** | **1.16** | **1.39** | **1.95** |
| | | CI | | **(ref)** | (1.05-1.28) | (1.21-1.59) | (1.46-2.62) |
| **Spine Fracture** | N (row %) | | 827 | 297 | 313 | 188 | 29 |
| | Unadjusted Rate (/1000 PYAR) | | 1.8 | 1.1 | 2.2 | 4.7 | 8.1 |
| | Fully Adjusted | RR | | **1.00** | **1.60** | **2.63** | **3.49** |
| | | CI | | **(ref)** | (1.34-1.89) | (2.13-3.25) | (2.31-5.25) |
| **Unspecified Fracture** | N (row %) | | 1,335 | 555 | 509 | 234 | 37 |
| | Unadjusted Rate (/1000 PYAR) | | 2.9 | 2.0 | 3.6 | 5.8 | 10.4 |
| | Fully Adjusted | RR | | **1.00** | **1.34** | **1.69** | **2.41** |
| | | CI | | **(ref)** | (1.17-1.52) | (1.42-2.01) | (1.68-3.44) |

**Supplementary Table 4: Fracture rate by eFI group**

Adjusted for age, sex, BMI, smoking, alcohol, osteoporosis, season and year of study

p<0.001 for all fracture sites by likelihood ratio test

PYAR= person-years at risk    RR=Rate Ratio    CI= 95% confidence Interval

| | | EFI Group | | | | Participants | Person-Years |
|---|---|---|---|---|---|---|---|
| | | Fit | Mild | Moderate | Severe | | |
| **Fully Adjusted** | RR<br>CI | 1.00 | 1.34<br>(1.26-1.43) | 1.68<br>(1.56-1.82) | 2.26<br>(1.93-2.65) | 113,779 | 466,923 |
| **with Ethnicity** | RR<br>CI | 1.00 | 1.28<br>(1.17-1.40) | 1.77<br>(1.57-1.99) | 2.44<br>(1.92-3.09) | 49,545 | 203,817 |
| **with IMD** | RR<br>CI | 1.00 | 1.31<br>(1.21-1.42) | 1.74<br>(1.57-1.94) | 2.34<br>(1.90-2.88) | 60,376 | 239,484 |
| **with bisphosphonates** | RR<br>CI | 1.00 | 1.37<br>(1.29-1.45) | 1.75<br>(1.62-1.90) | 2.38<br>(2.03-2.80) | 113,779 | 466,923 |

**Supplementary Table 5: Sensitivity Analyses on rate of any fracture by eFI group**
Additionally adjusted for age, sex, osteoporosis, season, year of study, smoking, alcohol and BMI

| | | | Total | Drug Class | | | P-Value |
|---|---|---|---|---|---|---|---|
| | | | | CCB | ACE/ARB | Thiazide | |
| **Any Fracture** | Fully Adjusted | N=<br>RR<br>CI | 6,567 | 2,790<br>1.00 | 2,003<br>0.99<br>(0.93-1.05) | 1,774<br>0.97<br>(0.91-1.04) | p=0.78 |
| **Hip Fracture** | Fully Adjusted | N=<br>RR<br>CI | 2,080 | 848<br>1.00 | 636<br>1.02<br>(0.92-1.14) | 596<br>0.98<br>(0.87-1.10) | p=0.78 |
| **Arm Fracture** | Fully Adjusted | N=<br>RR<br>CI | 2,325 | 1,019<br>1.00 | 677<br>0.90<br>(0.82-1.01) | 629<br>0.98<br>(0.88-1.09) | p=0.18 |
| **Spine Fracture** | Fully Adjusted | N=<br>RR<br>CI | 827 | 355<br>1.00 | 262<br>1.01<br>(0.85-1.19) | 210<br>1.01<br>(0.85-1.21) | p=0.99 |
| **Unspec Fracture** | Fully Adjusted | N=<br>RR<br>CI | 1,335 | 568<br>1.00 | 428<br>1.08<br>(0.95-1.24) | 339<br>0.95<br>(0.82-1.09) | p=0.19 |

**Supplementary Table 6: Rate of Fracture by Drug Class for specific fracture sites**
ACE/ARB= Angiotensin Converting Enzyme Inhibitor or Angiotensin receptor blocker, CCB= Calcium channel blocker
RR: Rate Ratio, CI= 95% Confidence interval. P-values from likelihood ratio test.
Adjusted for Age, sex, season, year of study, smoking, alcohol osteoporosis and BMI

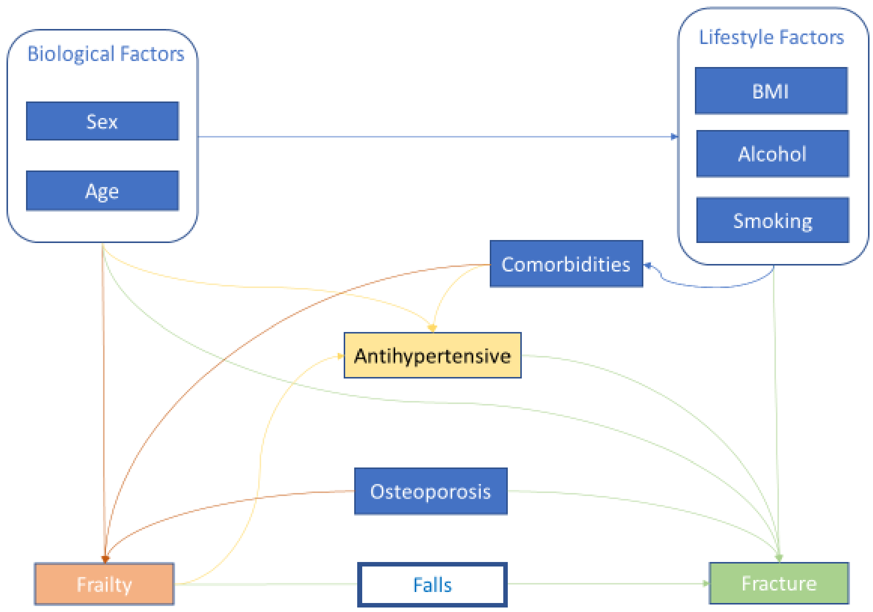

**Supplementary Figure 1:** Conceptual Framework
